# New Directions from the COMPASS Study: Participation and Communication in Rural Health Research

**DOI:** 10.1101/2025.03.04.25323155

**Authors:** Heidi Rishel Brakey, Maria-Eleni Roumelioti, Jesus E Fuentes, Darren W Schmidt, Larissa Myaskovsky, Christos Argyropoulos

**Affiliations:** University of New Mexico Clinical and Translational Science Center; Division of Nephrology, University of New Mexico

**Keywords:** Chronic kidney disease, kidney health, community research dissemination, plain language, research participation perceptions, results interpretation, National Kidney Foundation visualization tools

## Abstract

**Background:** The **Com**munity Based Study of the E**p**idemiology of Chronic Kidney Dise**as**e in Cuba New Mexico and **S**urrounding Areas (COMPASS) was designed to screen for chronic kidney disease (CKD) and discover novel related biomarkers in rural New Mexico, NM. As part of this study, we qualitatively explored participants’ opinions about CKD research and best practices for delivering lab results to patients.

**Methods:** This cross-sectional descriptive qualitative study was part of a larger longitudinal, epidemiological community-based mixed methods project. In COMPASS, participants were aged 18-80 years; lived in or near Cuba, NM; and had up to seven study visits over five years, including receiving a kidney lab results letter using National Kidney Foundation (NKF) visualization tools. All participants were invited to participate in an interview after one year, the focus of the current manuscript. We asked them about their thoughts of research participation and solicited feedback on the results letter. Using a team-based, iterative process, we elicited themes from transcribed interviews using NVivo software.

**Results:** We interviewed 33 adults of whom were 64% Hispanic, 24% American Indian, 55% female, 67% aged ≥50 years, and 42% high school graduates. Interviewees were positive toward participating in kidney health research; they appreciated the results letter, but most said they needed help interpreting and/or had suggestions for improvement. Many made positive lifestyle changes.

**Conclusions:** Community members in one rural NM area embraced the opportunity to participate in kidney health research. The NKF visualization tools were well-received and inspired positive lifestyle change, but results should be written in plain language. The letter demonstrates the potential efficacy of such interventions to improve understanding and care of medical conditions but also illustrates the opportunity to improve the effectiveness of this type of communication.

## Introduction

Chronic Kidney Disease (CKD) is a frequently unrecognized health condition associated with various comorbidities until end-stage kidney disease (ESKD) develops and initiation of maintenance dialysis becomes necessary. The distribution of CKD in the United States assumes a disproportionate importance in rural areas. While unique socioeconomic factors and limited access to both primary and specialist care play a considerable role in the disproportionate impact of CKD,^1–3^ additional factors cannot be ruled without CKD research programs that study rural areas to better understand the disease. Such programs will fulfill a secondary role: uncovering CKD or risk factors in study participants leading to therapeutic interventions slowing the rate of CKD progression and delay onset of dialysis, which are difficult to implement in rural areas.^4^

It is increasingly recognized that clinical practice and patient health outcomes can benefit significantly from research participation by healthcare providers and their patients.^5^ However, practicing in rural areas pose challenges to engage in research including lack of time or interest due to work overload and disruption of workflow.^6^ Meanwhile, researchers also face numerous challenges when striving for community engagement in research, such as understanding best practices of community-engaged research, the community’s culture, language, socioeconomics, structure/politics, potential mistrust toward research or medicine, and more.^7–11^ Regardless, community engagement is important to build trust and improve clinical and research outcomes.

New Mexico (NM) is unique in many ways: it has a rich culture with more than half its population being Hispanic or Native American; it is the fifth largest state in landmass, shares a border with Mexico, and approximately half of its population lives in rural areas.^12,13^ While these may be seen as strengths for research opportunities, it requires a special skillset of investigators for strong community engagement. Further, NM has challenges such as provider shortages, poverty, and transportation, particularly in rural areas, which may lead to additional barriers of in-person clinical research participation.^12,14^

To better understand whether our approach to community research was appropriate for the challenges in our area, we surveyed the participants in the study, ***<u>Com</u>****munity Based Study of the E**<u>p</u>**idemiology of Chronic Kidney Dise**<u>as</u>**e in Cuba NM and **<u>S</u>**urrounding Areas **(COMPASS).***^4,15^ This project was designed as a community-based kidney health screening and translational biomarker research program. An important secondary aim, and the focus of this manuscript, was to explore participants’ perspectives about CKD research and best practices for delivering kidney lab results to patients through qualitative interviews.

## Methods

This cross-sectional, descriptive, qualitative study was part of a larger, longitudinal, epidemiological community-based mixed methods project. Eligible participants were aged 18 to 80 years with a mailing address within 20 miles of the rural town of Cuba, NM. Patients were ineligible if they had a history of renal replacement therapy (dialysis or transplantation). We conducted interviews from April 2017 through December 2020, which included verbal informed consent separate from the overall COMPASS consent. Each interview lasted up to 30 minutes and was audio recorded and professionally transcribed, and participants received a $20 merchandise card. Prior to the COVID-19 pandemic, we held these interviews in-person at the dialysis clinic in Cuba; all interviews in 2020 occurred over the phone or zoom.

In the parent study, participants had up to seven study visits over five years that included questionnaires, clinical examinations, and blood and urine samples. They received a kidney lab results letter using the National Kidney Foundation visualization tools available at the time of study design (mid-2010s; Figure 1), and a follow-up letter with incidental lab results needing attention.^16^ All participants were invited to participate in an interview after their one-year visit, which is the focus of the current manuscript. This protocol is published,^4^ and this study was approved by the University of New Mexico Human Research Review Committee (HRRC #15-575).

**Figure 1.**
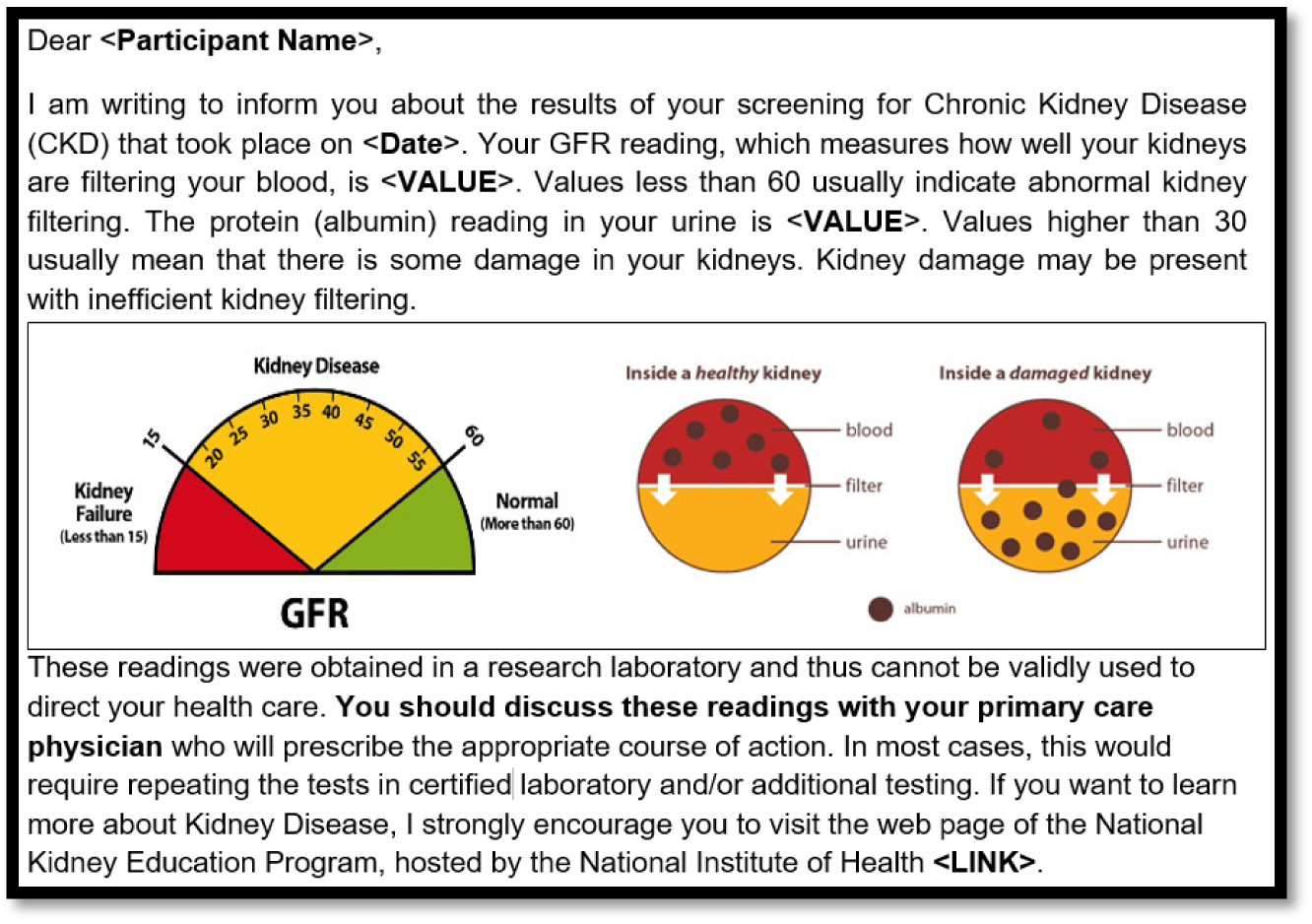
One kidney lab results letter template using National Kidney Foundation visualization tools^14^ for someone who has low GFR and high albumin. Letters also included contact information for the study PI.

The goal of these interviews was to understand participants’ thoughts about and experiences with the COMPASS study and research participation generally. We asked patients about their decision to join COMPASS, experience participating, health priorities, and suggestions for conducting research within their community. See Table 1 for interview guide.

**Table 1.**
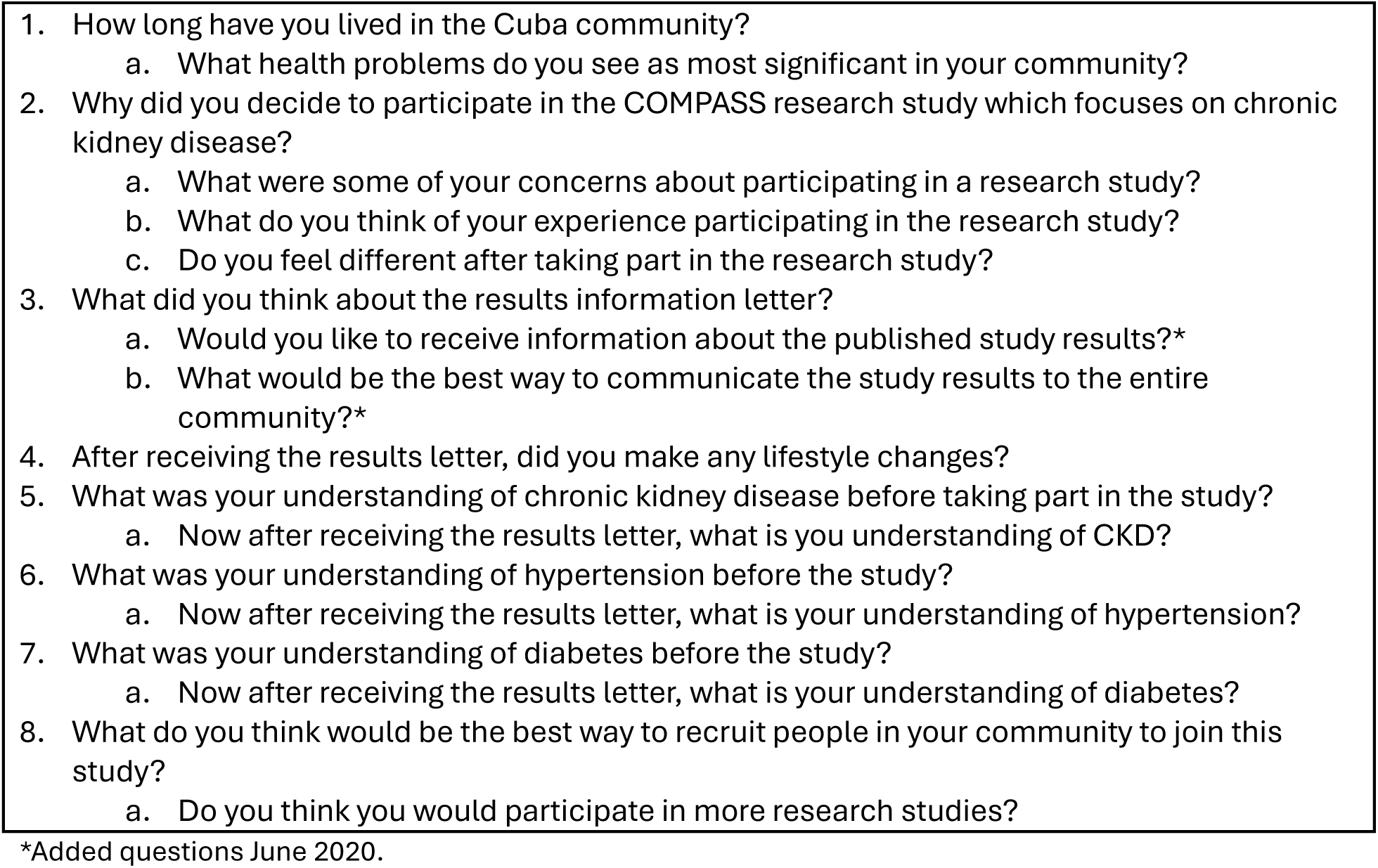
Primary questions of the semi-structured interview guide (not including prompts).

We conducted descriptive, qualitative analyses, using a team-based, iterative process and NVivo 13 (2020, R1, QSR International). The primary analyst JEF created a preliminary codebook deductively using the interview guide, inductively adding to it as new codes and themes emerged. He and the senior qualitative methodologist HRB independently coded five transcripts, meeting to discuss after each. Upon agreement, JEF coded the remaining transcripts. We conducted thematic analysis but used matrix queries to determine the number and percentage of participants with quotes in each code. We conducted content analysis only for responses to questions about chronic kidney disease, hypertension, and diabetes, to get an understanding of what participants learned from participating in COMPASS. Throughout, the analysts met regularly with the principal investigator CPA and content expert LM to discuss coding. As this project was descriptive, we did not begin with theory and our goal was not to reach saturation, but to hear from all participants in the study who were interested in an interview.

## Results

Of the 213 people who participated in at least one COMPASS visit, 118 were eligible to participate in an interview, of whom 33 accepted and completed (response rate=30%). Participants were 64% (n=21) Hispanic, 24% (n=8) American Indian, 54% (n=18) female, and 67% (n=22) aged 50 years or older. See Table 2 for demographics. The largest percentage of participants were high school graduates (n=12, 42%) and were employed (n=12, 36%) or self-employed (n=5, 15%).

**Table 2.**
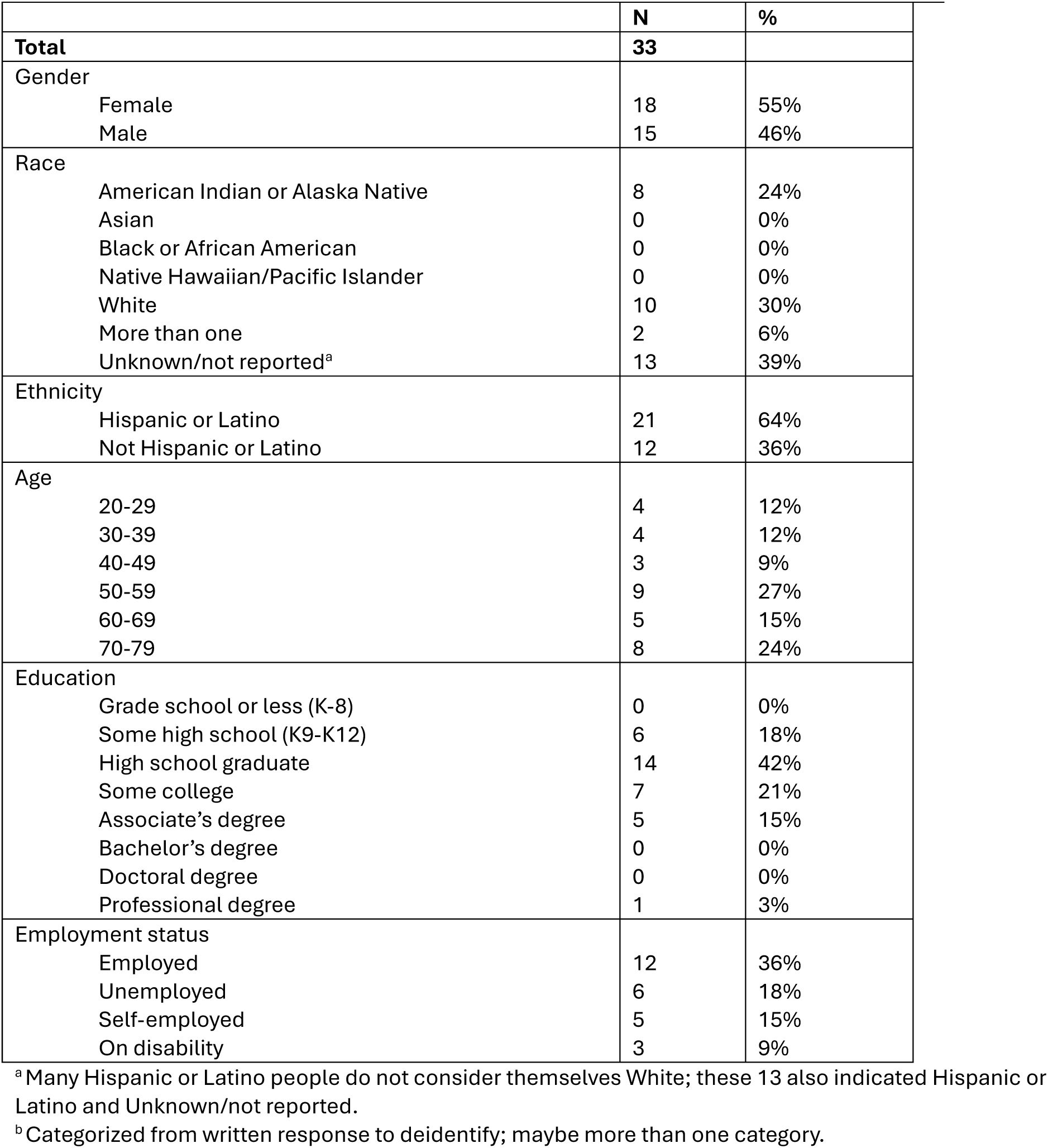
Self-reported demographics of interview participants at baseline.

|  | **N** | **%** |
| --- | --- | --- |
| **Total** | **33** |  |
| Gender |  |  |
| Female | 18 | 55% |
| Male | 15 | 46% |
| Race |  |  |
| American Indian or Alaska Native | 8 | 24% |
| Asian | 0 | 0% |
| Black or African American | 0 | 0% |
| Native Hawaiian/Pacific Islander | 0 | 0% |
| White | 10 | 30% |
| More than one | 2 | 6% |
| Unknown/not reported<sup>a</sup> | 13 | 39% |
| Ethnicity |  |  |
| Hispanic or Latino | 21 | 64% |
| Not Hispanic or Latino | 12 | 36% |
| Age |  |  |
| 20-29 | 4 | 12% |
| 30-39 | 4 | 12% |
| 40-49 | 3 | 9% |
| 50-59 | 9 | 27% |
| 60-69 | 5 | 15% |
| 70-79 | 8 | 24% |
| Education |  |  |
| Grade school or less (K-8) | 0 | 0% |
| Some high school (K9-K12) | 6 | 18% |
| High school graduate | 14 | 42% |
| Some college | 7 | 21% |
| Associate’s degree | 5 | 15% |
| Bachelor’s degree | 0 | 0% |
| Doctoral degree | 0 | 0% |
| Professional degree | 1 | 3% |
| Employment status |  |  |
| Employed | 12 | 36% |
| Unemployed | 6 | 18% |
| Self-employed | 5 | 15% |
| On disability | 3 | 9% |

<sup>a</sup> Many Hispanic or Latino people do not consider themselves White; these 13 also indicated Hispanic or Latino and Unknown/not reported.
<sup>b</sup> Categorized from written response to deidentify; maybe more than one category.

We identified two primary themes: “Community” and “COMPASS.” “Community” included codes of health problems and resources. “COMPASS” covered the spectrum of participant involvement in a study: decision-making about participating; experiences with participation; health education they received during the study; recommendations to improve study experience; and interest in dissemination and potential future research participation. See Table 3 for codebook, including example quotes and the number and percentage of participants who spoke about each code. See Appendix 1 for an expanded codebook with additional quotes. We now consider these themes in greater detail.

**Table 3.**
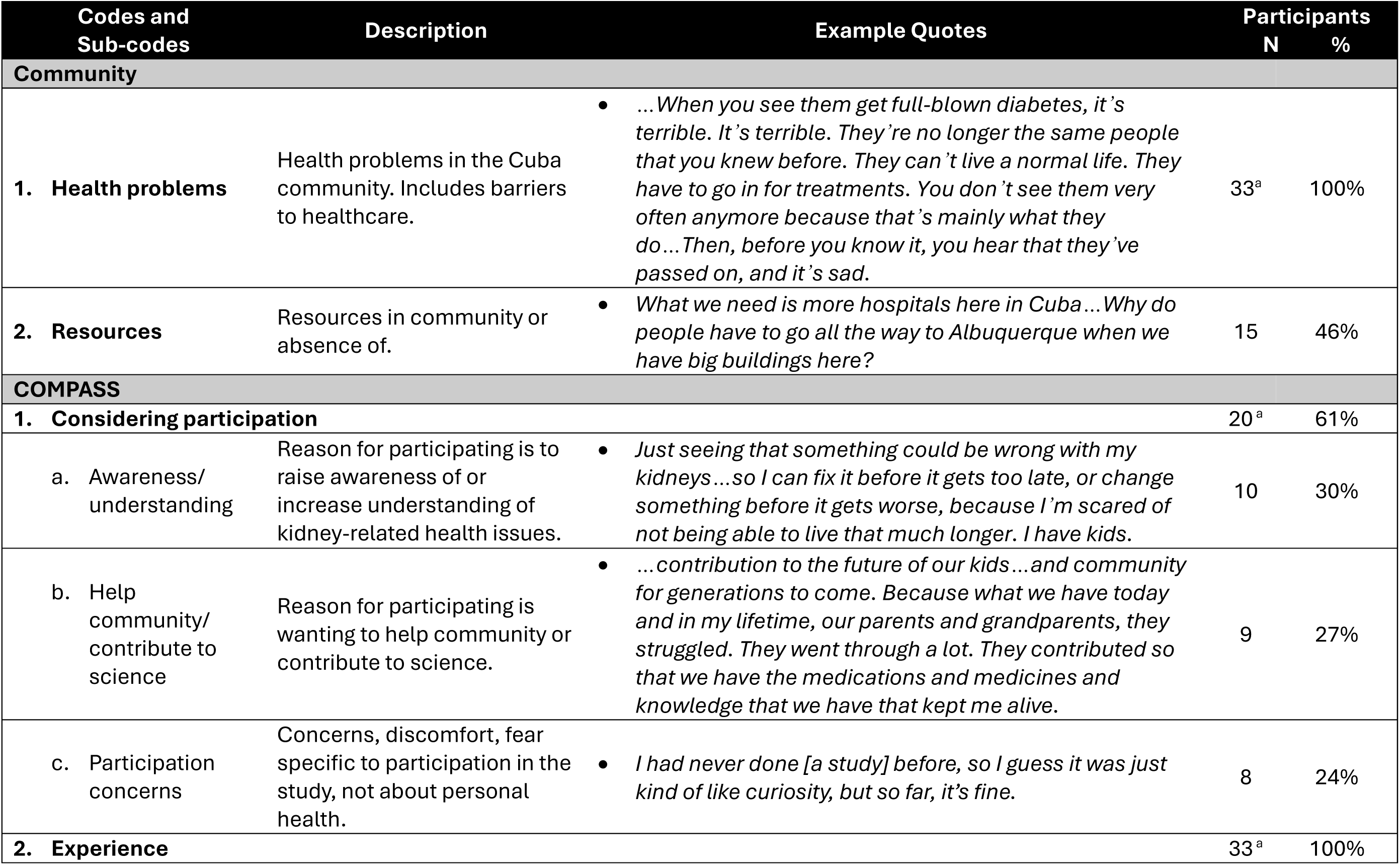
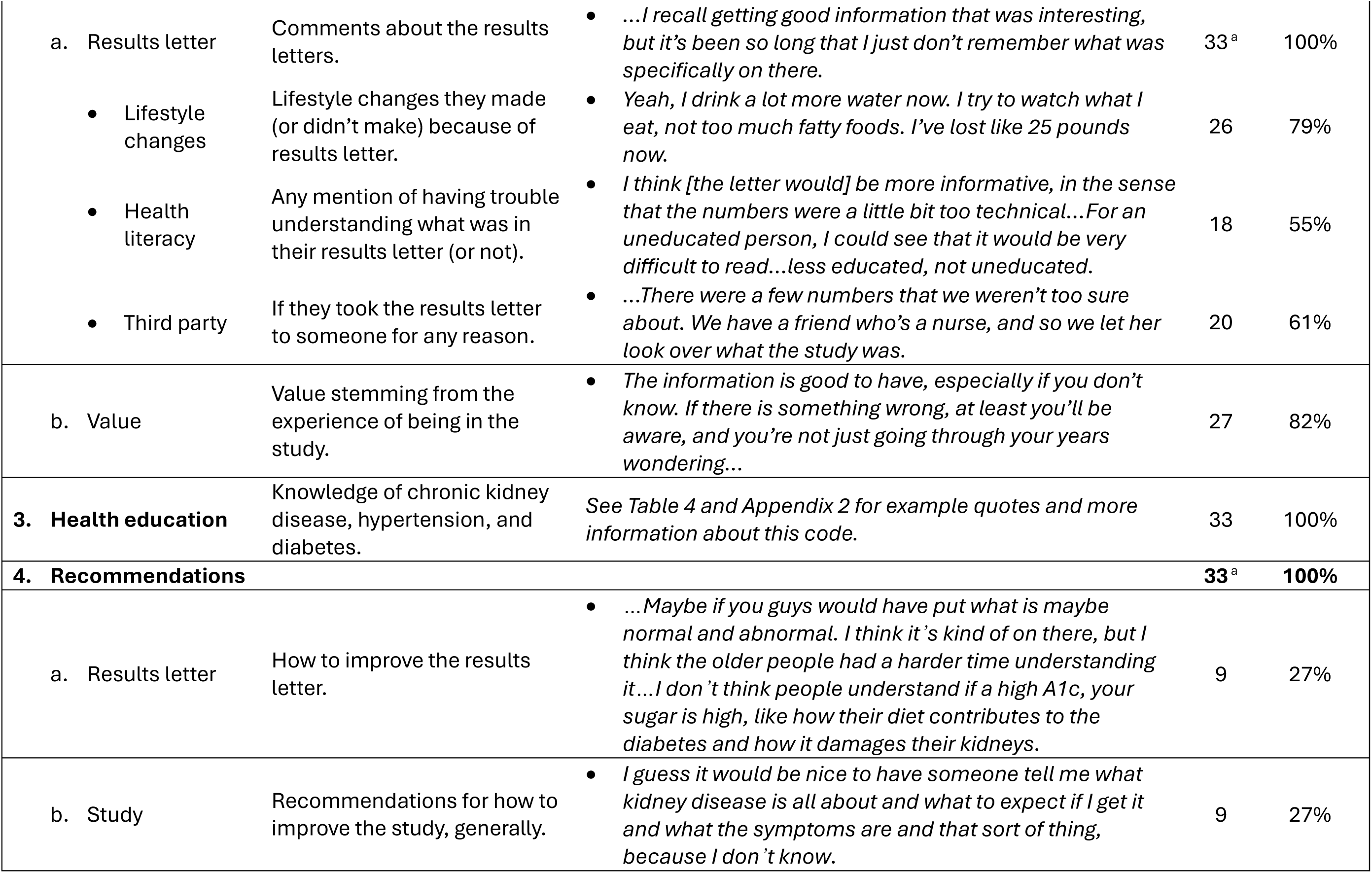
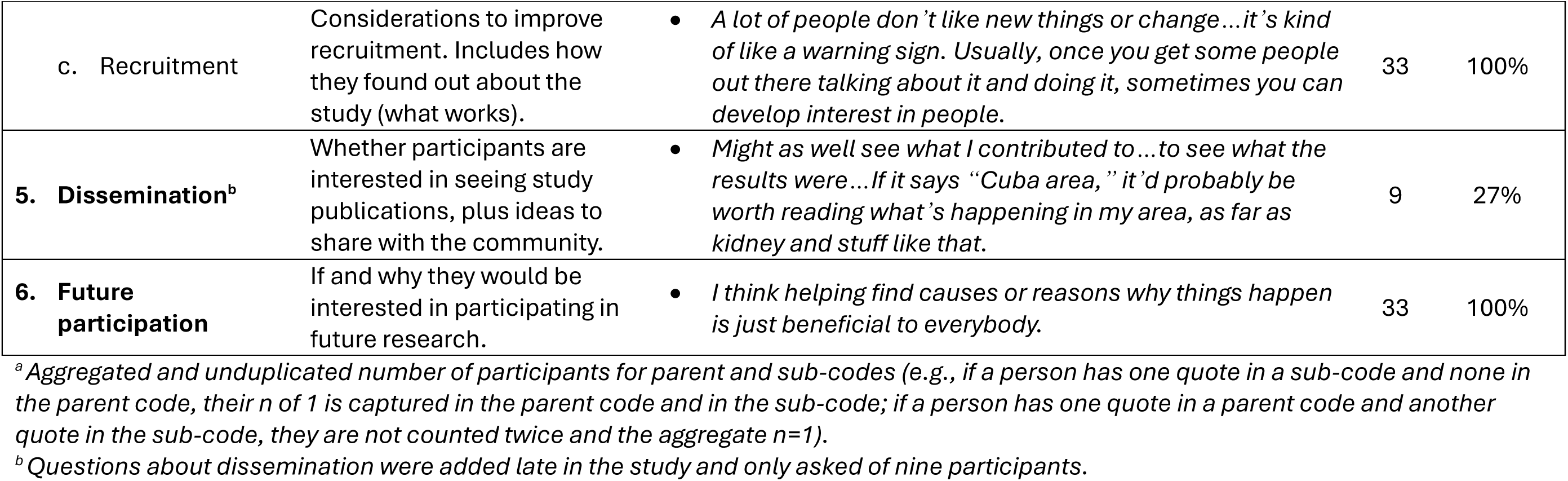
Codebook, including example quotes and number and percentage of participants with quotes related to each code, split between main themes of Community and COMPASS.

### Community

Participants spoke about health problems and resources in and around Cuba (see Table 3 for example quotes). They frequently cited diabetes and chronic kidney disease as the most significant health problems. Many had experience or knew someone with these conditions. Several suggested reasons for this, including lack of exercise, poor diet, or low income making it difficult to purchase healthy foods. Other health problems included dementia, Alzheimer’s, arthritis, cancer, hypertension, methamphetamine use, and alcohol use. They attributed some of these issues to lack of resources in their community. For example, many people must travel at least an hour for healthcare. Others mentioned poor water quality, and being a rural area, there are few options for fresh food.

### COMPASS

Most of the discussion revolved around the COMPASS study. This included considering participation, their experience participating, health education, recommendations to improve studies like COMPASS, dissemination ideas and preferences, and whether they would consider future participation in another study. See Table 3 for example quotes.

#### Considering participation

When asked why they decided to participate in COMPASS, participants discussed two primary reasons: to raise awareness of or increase their understanding of kidney-related health issues; and to help their community or contribute to science. For awareness and understanding, one person discussed putting *“light on such a subject”* and another wanted to know if she needed to fix her kidneys *“before it gets too late.”* To help the community, one participant said it was his *“contribution to the future of our kids.”* Others cited the need to contribute to science, as one woman said, “*Because I believe in research studies.”*

We asked if they had concerns about participating in research. Most said they were not concerned prior to participation, but a few said they had wanted more information about what participation would mean, including what they would get from it and what they needed to do. Two women were nervous because one was “*not used to going to the doctor,”* and another did not like needles.

#### Experience

Participants had much to say about their experience in the study, specifically commenting on the results letter, subsequent lifestyle changes, and any value they gained. When asked about the letter, several people had difficulty remembering much, if any, of its contents since they received it a year previously. Most people had a general idea of what it said, however, found it valuable, and many made lifestyle changes as a result. One man said it opened his eyes and, *“It made me aware that, really, what I put into my body is what I’m getting out of it, in terms of alcohol or junk food, and in terms of diabetes and, of course, liver damage…That really did open my eyes to a lot of that stuff, but I thought…it didn’t apply to me, but I guess I was just fooling myself.”*

Some said the letter made them want to be healthier, generally; others said they started eating *“a lot of greens,” “trying to eat less fatty foods,”* and *“drinking more water daily.”* Two said they decreased alcohol consumption, one stopped taking as much ibuprofen, and a few began exercising more. One person said they even lost 25 pounds because of diet changes as a result of the letter. Conversely, some people said they did not make many changes, if there were any, because they were not interested or because the letter said they were healthy. Two said they tried to be healthier and then *“lost interest”* or stopped trying as much, partially because of external factors, as one man described: “*I tried to drink more water and not sod…after a while, it just kind of went by the wayside…”*

Some participants said the letter was easy to understand, but many said they had trouble understanding it, that it was *“too technical” or “very difficult to read.”* Many asked providers, family, or friends to help interpret their results. One person took it to his nurse friend, and another found more information online. Aside from interpretation, some people went to their providers for a second opinion or as a follow-up to the letter.

Many participants said their experience participating in the study was valuable, namely because of the information in the results letter. Two people said there was value in getting screened because they had not received this information from a healthcare provider. Some said it was good to know whether they were healthy, while others said it was good to know they needed to make changes.

#### Health education

We asked participants about their understanding of chronic kidney disease, hypertension, and diabetes before they joined the study and then after they received the results letter. Table 4 shows the number of participants who had personal experience or understanding, whether they knew more about these health topics after participating in the study, and example quotes. See Appendix 1 for all quotes coded to each category.

**Table 4.** Participants’ response to the prompts, “What was your understanding of [chronic kidney disease, hypertension/high blood pressure, or diabetes] before taking part in the study?” (pre) and “Now after receiving the results letter, what is you understanding of CKD [chronic kidney disease, hypertension/high blood pressure, or diabetes]?” (post). Includes example quotes and number of participants for each type of response. See Appendix 2 for all quotes and their corresponding codes.

|  | Chronic Kidney Disease<br>n (%) | Hypertension<br>n (%) | Diabetes<br>n (%) |
| --- | --- | --- | --- |
| Total | 33 | 33 | 33 |
| Had personal* experience or understanding (pre) | 12 (36.4) | 7 (21.2) | 16 (48.5) |
| Maybe knows more (post) | 7 (21.2) | 8 (24.2) | 6 (18.2) |
| Knows more or has decent understanding (post) | 10 (30.3) | 8 (24.2) | 8 (24.2) |
| Does not know more (post) | 16 (48.5) | 17 (51.5) | 19 (57.6) |
| **Example pre- and post-quotes** |  |  |  |
| Chronic Kidney Disease |  |  |  |
| <u>Pre</u> (personal experience): *"My understanding of kidney disease is what's been in my family for as long as I can remember. It has killed numerous people in my family. It has taken their life, and it's scary."*<br><u>Post</u> (does not know more): *"I'll be honest with you. I mean, our history goes back so long that, what is there to learn? It's hereditary for us, and it's a bad disease."*<br><u>Pre</u>: *"Not a whole lot. I didn't know much, the functions of it...I think I [had heard of it], but I didn't pay much attention to it because I've never had an issue with it or didn't know anybody with issues with their kidneys."*<br><u>Post</u> (knows more): *"I think it's affected by medications and things that you take, and even food you eat and the amount of the water you drink. It all ties into healthy kidneys."* |  |  |  |
| Hypertension |  |  |  |
| <u>Pre and post</u> (do not know more): *"I don't know anything about high blood pressure. I don't know how to answer that."*<br><u>Pre</u>: *"Bad eating, weight, diabetes causes high blood pressure, and it's not good."*<br><u>Post</u> (maybe knows more): *"Same understanding. Just what to do about it now...You take medication, exercise, all that stuff that I'm not doing. Eat better. Lower salt. I don't know if that's a thing."* |  |  |  |
| Diabetes |  |  |  |
| <u>Pre</u>: *"Sugar diabetes? Well, my understanding of diabetes is that there's a problem with the blood sugar level, which we can get at any time, especially as we get older, so it's just good to participate."*<br><u>Post</u> (maybe knows more): *"I think it's all good. We have a pretty good idea. My [spouse] likes keeping up with the stuff, so [they keep] me posted."*<br><u>Pre</u> (personal experience): *"Diabetes was when you had a lot of sugar. That's the way I thought it was..."*<br><u>Post</u> (knows more): *"That's the way I thought it was, when you eat too much sugar, but it's not. It has a lot to do with your triglycerides, your—all these other fancy words that I can't say, because I've learned all those, believe me...I'm learning, because I just got diagnosed when I was fatter, and now that I'm losing it, my numbers are coming down. I'm learning about diabetes at the same time."* |  |  |  |

\*Self, family, friends, or professional

Many mentioned previous experiences with chronic kidney disease (n=12, 36.4%), hypertension (n=7, 21.2%), and diabetes (n=16, 48.5%); because of this, some said they did not gain knowledge from the letter. Experience did not mean they knew much about the disease prior to COMPASS, however, and some still gained knowledge, as one man described:

Before participating: *“…Because my mother had [CKD], and my grandma has it, and my auntie has it, and my uncle has it, I just knew that they would just go get connected to a machine and that you really couldn’t pee no more. That’s all I pretty much knew.”*

After participating: *“Now I’ve learned that it can run in the family, also that if you don’t take care of your diabetes, you could get it…I thought you just got low blood sugar when you weren’t eating right on diabetes. I didn’t know you could lose limbs or get kidney failure and have to go on dialysis.”*

Regardless of experience or knowledge, most (n=28, 76%) did not know more about at least one of the three diseases after participating. Indeed, many did not even know what we were asking, particularly with the word “hypertension,” as exemplified in the following exchange:

Interviewer: “What was your understanding of hypertension before the study?” Participant*: “I don’t know. Hypertension would be like you’re very hyper and stuff like that. You get nervous, all that stuff.”*

Interviewer: “After receiving the results letter, what’s your understanding now?” Participant*: “How do I understand it?”*

Interviewer: “Yeah, I’m just trying to ask about how your understanding might have changed before and after the study.”

Participant*: “Yeah, it would change. I would understand more what it’s about, how to go about it and everything, how to help myself, somebody like that can work with me or something.”*

#### Recommendations

We asked participants for recommendations to improve the results letter and the study overall, and how to recruit in their community. The most common recommendation to improve the letter was to write it in more plain language with explanations of tests and implications in a way that more people could understand. One woman said she thought the letter was a *“scare tactic”*:

*…It was making it sound like I had a kidney disease and I needed to talk to my provider immediately…He did check me out. He did find out there was nothing wrong with me…It was not as informative as I think it probably could have been. In other words, saying that, ‘We did find this in your kidney. You might want to talk to your provider, but you don’t have a kidney disease,’…To me, it felt like a form letter. Like, whoever was doing this project, we’re trying to cover their butt as far as making sure that they don’t get sued for some crazy stuff. I understand that…but still it was like a scare tactic because it scared the crap out of me. I thought I had a kidney disease.”*

Others said they wanted more details about the various tests in the letter and one person would have liked to receive the letter sooner after their study visit.

Participants suggested other ways to improve health education, such as holding community classes, providing more written information, and follow-ups in person or over the phone. Everyone gave recommendations to improve recruitment or discussed how they learned about the study as a method that works for their community. Many learned about the study word of mouth and also told others they knew about the project. Flyers were widely cited as a viable recruitment method and suggested other venues such as Craigslist, the newspaper, or tabling at the post office, health fairs, the senior center, and special events.

Some said it is important to build trust by spending time with and *“getting more involved in the community.”* A couple of people suggested doing this by holding education classes or, as one woman said,

*“Just make a presentation somewhere; tell them that you’re not going to hurt them. A lot of people in this area…they’re scared…you have to really convince them that it’s okay, you’re not going to scare them, it’s not going to hurt, because they’re set in their ways, and their ways are the old ways.”*

Finally, adding flexibility could increase participation. For example, expanding study inclusion to neighbouring counties and having visits available evenings and weekends.

#### Dissemination

Of those we asked if they would be interested in receiving published study results, but one said they were. When we asked for suggestions about community dissemination, they recommended sharing through social media, bulletin boards, the Cuba clinic, direct mail, or presenting it as a seminar.

#### Future participation

Finally, everyone said they would or may be interested in participating in future research. Only four people said they would participate, depending on the specifics of the study (e.g., “the whole needle thing”), including scheduling and time commitment. When asked why they would be interested, their reasons were similar to what they gave for participating in COMPASS: most were interested in learning about their own health and how to prevent or treat disease; a couple cited earning money; others said they believe in research, that it is beneficial to everyone, and they want to help their community.

## Discussion

This unique report provides insights from a subset of participants in the COMPASS study. Overall, rural community members in Cuba, NM, embraced the opportunity to participate in kidney health research with positivity and enthusiasm. The visualization tools for displaying lab results were well-received, but patients could use help interpreting results. The results letter, however, still inspired positive lifestyle changes for many, and underscores the beneficial impact of clinical research in these communities. Herein we discuss some major takeaways from our discussions with recommendations and suggestions for future studies.

Regarding reasons for research participation, we found our study participants thought collectively about community members, family, and friends in addition to increasing their understanding of kidney-related health issues. They wanted to raise awareness and hence help their community and contribute to science and to the future of their kids. When asked, they also provided some concerns about participation in a study: some were concerned about potential benefits while others expressed concerns about the medical visit process. These concerns underscore the importance of providing clear and informed consent to participants with concise and easy-to-understand information about potential benefits and risks. We should also provide, if feasible, alternative resources for health education and health provision. Finally, when asked how they heard about the study many responded through word of mouth and provided useful recommendations on how to improve study recruitment.

Participants discussed many important points related to the results letter. Most found it useful, valuable, and mentioned important lifestyle changes they made as a result, like exercising more, drinking less alcohol and more water, improving dietary habits, and losing weight. Of note, some participants said this was the first time they had been screened for kidney function. Even with these positive outcomes, many participants had difficulty understanding the letter and reached out to physicians, family, friends, or the Internet for help. We suggest the use of community advisors or community health workers to help write letters in plain language and be available for interpretation.

The code of health education included participants’ understanding of CKD, hypertension, and diabetes before joining the study and after receiving the results letter. Interestingly, approximately half of participants did not gain knowledge about these topics. Particularly, some people had trouble with the word hypertension but understood “high blood pressure.” This highlights the need to use plain language medical terms not only in the letter, but also while speaking with people in interviews or clinical visits. Some people expressed nervousness about the health education questions and felt like they “needed” to know the answers. This highlights the issue of potential power dynamics as well as the importance of using best practices in interviewing techniques (e.g., not asking “why” questions and putting people at ease).^17–19^ It was also clear that there is an unmet need for better health education and greater health literacy since misconceptions regarding the potential risk factors for CKD were unmasked from these interviews.

Our study seemed to benefit this rural community and posed a few limitations as well. Interviews occurred about a year after participants got the results letter from their first visit, so it was difficult for people to remember the content, what they thought about it, and/or if they made any lifestyle changes as a result. Participants also said diabetes/CKD is a health priority, but this could have been influenced by the questions in our interviews. To gain less biased answers, it would have been better to ask the community about their health priorities prior to the study. Finally, it was unclear whether participants actually gained knowledge about health conditions from their results letters. The incorporation of a pre- and post-study survey may help future studies more clearly define these gains. Extrapolating from this observation, we feel that future community studies or initiatives to increase awareness of CKD should integrate kidney related themes (e.g. Cardiovascular Kidney Metabolic syndrome).^20^

## Disclosures

We have no disclosures to report.

## Funding

The COMPASS Study is supported by grants from DCI & NCATS (UL1TR001449).

## Data Availability

All data produced in the present study are available upon reasonable request to the authors.

## Acknowledgements

This was part of a larger multi-year mixed methods study, and its success would not have been possible without the numerous individuals involved. We would like to thank all the participants of this study for their valuable time and input. We also would like to thank the various university experts involved, consultants, the CTSC research coordinators as well as staff at Dialysis Clinic, Inc for their assistance and for providing us with space to conduct this project.

## Author contributions

The research team was made up of nephrologists, kidney researchers, and content experts at the University of New Mexico’s Division of Nephrology and community engagement and qualitative research specialists at the University of New Mexico’s Clinical and Translational Science Center. All authors were involved in reviewing and editing drafts of the manuscript, interpretation of the final analyses, edited and approved the final version of the manuscript. HRB trained and oversaw the qualitative research specialists who conducted interviews and analysis; she reviewed and contributed to all analyses and wrote the methods and results section of this manuscript. MR wrote and mailed participant results letters, reviewed final analyses, and co-wrote the introduction and discussion of this manuscript. JEF conducted many of the interviews and preliminary qualitative analysis. DS provided clinical context for the interpretation of the analyses. LM provided content expertise and scientific oversight of the qualitative activities and the analyses of the project. CA was the project leader who designed the COMPASS protocol, obtained funding, and developed the qualitative and quantitative instruments used in the project.

## Data sharing statement

The Vivli (www.vivli.org) Center for Global Clinical Research Data will be used to generate a unique DataCite DOI for sharing of data and meta-data upon final acceptance for peer-reviewed publication.

## Acronyms

CKD: Chronic kidney disease
COMPASS: Community Based Study of the Epidemiology of Chronic Kidney Disease in Cuba New Mexico and Surrounding Areas
ESKD: End-stage kidney disease
NKF: National Kidney Foundation
NM: New Mexico

## Notes

### Competing Interest Statement

The authors have declared no competing interest.

### Author Declarations

The Human Research Review Committee (ethics committee/IRB) of the University of New Mexico Health Sciences Center gave ethical approval for this work.

