## Appendix 2 for "New Directions from the COMPASS Study: Participation and Communication in Rural Health Research"

**Appendix 2.** Participants' response to the prompts, "What was your understanding of [chronic kidney disease, hypertension/high blood pressure, or diabetes] before taking part in the study?" (pre) and "Now after receiving the results letter, what is you understanding of CKD [chronic kidney disease, hypertension/high blood pressure, or diabetes]?" (post).

| Pt* | **Chronic Kidney Disease** *(What was your understanding of chronic kidney disease before/after this project?)* | |
|---|---|---|
| | **Pre** | **Post** |
| 2 | I just knew that—because my [parent] had it, and my [relative] has it, and my [other relative] has it, and my [other relative] has it, I just knew that they would just go get connected to a machine and that you really couldn't pee no more. That's all I pretty much knew. | Now I've learned that it can run in the family, also that if you don't take care of your diabetes, you could get it. I also learned that you could lose body limbs from diabetes. I didn't know that. I thought you just got low blood sugar when you weren't eating right on diabetes. I didn't know you could lose limbs or get kidney failure and have to go on dialysis. |
| 3 | "Chronic" means serious. "Kidney disease"—we all realize that our water isn't the greatest. Your kidneys filter the water and the blood, so it's always good to find out to make sure everything's working properly. | ...it's improved...Probably just because we asked more questions, because we get a copy of the information when we come in, and so we go home and reread it and stuff to better understand what's going on, I guess. |
| 4 | Really, I don't know that much about it. | Now that I know and I've been asking around, it's really—how would you say—it's a problem. Especially out here, I hear a lot of different ones that are on dialysis and stuff, and it's something that a person would never really want to have to experience, so to do what you can to not be able to wind up having to get it. |
| 6 | I have always only worked with renal patients, so I was pretty familiar with the disease process. | Like I said, it's always been something that I've dealt with, with people I work with, also with family. So it's…nothing new. |
| 7 | I had somewhat of an understanding of it because of my family history. | I would say no. The results letter does not really, I guess, educate somebody on it. |
| 14 | My understanding of kidney disease is what's been in my family for as long as I can remember. It has killed numerous people in my family. It has taken their life, and it's scary. | I'll be honest with you. I mean, our history goes back so long that, what is there to learn? It's hereditary for us, and it's a bad disease. |
| 16 | I took it lightly, I guess, because I wasn't aware of all that really takes place with all that. Now, I can kind of see the seriousness of it. | I would say both, of being in the study and what takes place, all that you guys have to do to check it out, but then when I got the results as well, it was kind of comforting. |
| 27 | I had, I think, a fairly decent idea about what it is. If the kidney output was—I guess it would be input, too, and output where it was at. If it was okay, if there was something else wrong, because sometimes even though I had yearly physicals, the physicals here in New Mexico compared to in [another state] are totally different. | Like anything else, early detection is key if you have something like that, because if you wait, i.e., my eyes, which I didn't know—I just thought it was whatever, it was like, age; maybe I'm just not seeing as good and it's time for another pair of glasses—it's the same kind of thing that if you know from the beginning, it's much easier and treatable kidney disease than it would be if you came already with symptoms, you weren't passing urine, blah, blah, blah, all of that. |
| 35 | Kidney disease is obviously pretty serious. Kidneys are very important. It's just like any filter system. If a filter doesn't work, then there is no filter. We need to have things removed from our blood just like everything else. You take in food and air and it needs to go through the system to work right for your health. | No, I'm quite aware of the biological processes of the human body. |
| 39 | I really didn't know. Like I said, all I heard was it runs strong. I didn't know until after I did this test, but now I understand a lot. | I understand what causes it and what to do—well, not what causes it, because I'm not a doctor, but what to do and how to avoid certain things. |

*Participant ID is unknown to everyone outside the research team.

Key:

| Had personal experience or understanding (pre) | Maybe knows more (post) | Knows more or has decent understanding (post) | Doesn't know more (post) |
|---|---|---|---|

| Pt* | Chronic Kidney Disease<br>*(What was your understanding of chronic kidney disease before/after this project?)* | |
|---|---|---|
| | **Pre** | **Post** |
| 41 | My [sibling] has polycystic disease. [They] also had kidney stones. [They] had been treated most of [their] adult life for these conditions, and so from [their] experience would be what I know…[they] had a kidney transplant also. | I don't think it's influenced [my understanding]. |
| 54 | I understood a little bit of it, because I know some of the people that come to this dialysis center, and I know it's not a—it takes a really bad toll on a lot of people, but as far as before, I really never really thought about it, to tell you the truth, but after it was like, "Wow, yeah," you start think, and then you get busy. | Oh, definitely [it helped understanding]. I can let my elders know that this program is available, and if they're feeling like they really need to—and not just the elders, anybody, anyone that feels like they need to go and have themselves checked; I'll send them over here. |
| 57 | No, I don't know. | here's nothing wrong with my kidneys, so I guess there's no disease. I don't have very much to say about nothing. All I know is that I'm okay...I don't know nothing [about kidney disease]. I'm not into medication or anything like that. Doctors. I just live a simple life. |
| 67 | Kidney disease—for some reason, here in Cuba, I don't know if it's the water, if it's the environment; something is going on, and I figured with my other health problems, see how that works in with everything else, as far as diabetes and kidney disease and all of that. | Not really [understanding didn't change]. Just, it's a problem for everybody. |
| 68 | I knew quite a bit of it because of family. | There were a few things that I wasn't aware of. I mean, you got to take care of your body to eliminate some of the problems. Some of the problems can be taken care of by management. |
| 71 | Pretty much didn't know nothing about it. | A little more, not that much more. I couldn't name stuff off right now because, like I said, I read that letter, except I can't remember what it was...If you'd have called me right after I received the letter, I could probably tell you everything you need to know, but like I said, it's been a while. |
| 76 | Just that if you got kidney failure and that you'd probably have to go to dialysis. That's pretty much all I knew. | Pretty much the same, I think. |
| 90 | I've taken care of people who were on dialysis, and they did not survive. You need your kidneys to live pretty much. There's just no doubt about that. It's dangerous when you don't understand how a person gets to that point, but it's important to know what to do to prevent that from happening in the first place, because people are very uneducated. They don't know; they just live like me. I just lived. I didn't know. I didn't read books in that field to understand. | I think if you just want to keep living recklessly, you will. I think that's a person's choice if they wanted to go that route, but there's consequences if there's no changes. I am not a very disciplined person. I would love to be more disciplined. I truly would. I really, truly would, but I'm [in my 50s], and I think that some people live to be 100. I don't think I'm going to live to be 100. I would love to live to be 100, but I don't think I am. It's like any kind of abuse, whatever abuse you do to your body with food, it's abuse. If you've lived that for [50-something] years now and you're like, "How am I going to fix that? How much more life, if I do fix it, will I have?" That's where I am at. |
| 110 | Chronic kidney disease, I would relate that to, that's a lot of boozing. And the water here, at times, when I was growing up in the '80s, looked like iced tea—sometimes really dark, the minerals that were passed through the well, the Cuba water that was being distributed around the community itself...I'm pretty | Yes, I see it as a very big problem. I know that it's fatal, if not the treated or acknowledged and just left as is. But I think to catch it at an early time is paramount if you expect to live a little longer and not be in need of dialysis. |

*Participant ID is unknown to everyone outside the research team.

Key:

| Had personal experience or understanding (pre) | Maybe knows more (post) | Knows more or has decent understanding (post) | Doesn't know more (post) |
|---|---|---|---|

| Pt* | Chronic Kidney Disease<br>*(What was your understanding of chronic kidney disease before/after this project?)* | |
|---|---|---|
| | **Pre** | **Post** |
| | sure that I put my kidneys through a heck of a lot, rather than just with booze. | |
| 116 | Not good [understanding]. | If I learned anything, we'd have to read about it instead of learn through the book or whatever, if I do look at it, which I don't. I have to go to the library and look everything up. I want to. |
| 117 | I don't have much of an understanding, other than the kidneys not functioning and filtering out the way it's supposed to. That's about all the understanding I have. The reasons behind it, I don't really—I know diabetes causes kidney failure. | I know a lot more things cause kidney failure now. It can be—well, now I say that, and then I don't know, but diabetes, for sure. Some people are hereditary—things like that, because I know somebody that their family there's a whole line of them that are doing that, just that it's not just from diabetes or anything like that, that it can be inherited from your family. |
| 119 | I know people that have it really severe have to have dialysis. I'm not real sure. I guess I don't really know too well what kidney disease is all about. I just know if they fail, you're in trouble. | I'm not sure. I don't think anything's changed. I really don't know. I guess I need to look it up and find out what it's all about and what the symptoms are, because I don't even know what those are. |
| 123 | My understanding of kidney disease is it's bad. That's it. | My understanding of kidney disease is it's bad. That's it. |
| 124 | I didn't understand it until, like I said, my kidney started failing and all that., but disease? I don't have no disease, other than just the idea that I don't know why my kidney went down. They did something. I don't know what it is, but it's going up now. | I understand that you've got to stay up to get examined and find out what's going on. That's about it. |
| 127 | It's about the same as what I know now. I didn't really gain much more information. | It's about the same as what I know now. I didn't really gain much more information. |
| 131 | I didn't know anything. I wasn't into that until I participated. Before, I had no clue about anything. | It's important now to make sure I keep doing good on my kidneys, because I know it's not easy to get a kidney if something happens. |
| 132 | I didn't really have a good understanding. The only thing I learned about it was when I had kidney stones, so I don't know. | I don't know, sorry. |
| 141 | Actually I've been a caregiver with a kidney transplant patient, so I very much know about the functions and everything about the kidneys. The patient that I work with had a lot of [their] health based on this kidney transplant he had. I pretty much—pre-care, post-care, and most of it, I understood. | Kidney disease, actually, it's not very good for me, for anyone, actually. It's just like it's something to really—it's very important. You don't want to go take that awful route down, like I said, the patient that I saw go through. |
| 145 | Not really anything. | I don't know. |
| 157 | At first I wasn't really too familiar with any kidney diseases or anything. The only thing I've heard about was just little things like dialysis. I had a family member that was dealing with kidney problems, so he used to go to dialysis. That's the only thing I knew about it. I didn't really know much or a lot about it. | Now I think it's a better understanding for my health, just in case my body were to ever go the route of kidney disease or anything like that. There's a research study that you could do that could help me prevent that or can help me better my health or something like that. |
| 166 | What's that, chronic disease? Never heard that before. That's my very first time I heard that...<br>Because I have a [sibling] that has a kidney problem, and [they're] the one that explains everything, what's going on with [their] kidneys. I don't know. Other than that, I got to hang around with [them]. I go with [them] to the place there in Albuquerque. When [they] took that stuff with [them], [they] was just feeling groggy and throwing up a lot. Other than that, I don't know. | I don't know. I think it's still the same. |

*Participant ID is unknown to everyone outside the research team.

Key:

| Had personal experience or understanding (pre) | Maybe knows more (post) | Knows more or has decent understanding (post) | Doesn't know more (post) |
|---|---|---|---|

| Pt* | Chronic Kidney Disease<br>*(What was your understanding of chronic kidney disease before/after this project?)* | |
|---|---|---|
| | **Pre** | **Post** |
| | That's why it's hard to say. I don't get along with none of my family now. | |
| 194 | Not a whole lot. I didn't know much, the functions of it...I think I [had heard of it], but I didn't pay much attention to it because I've never had an issue with it or didn't know anybody with issues with their kidneys. | I think it's affected by medications and things that you take, and even food you eat and the amount of the water you drink. It all ties into healthy kidneys. What I really didn't quite understand is how some people donate kidneys and can just live with one. |
| 195 | It's very devastating and that it runs in our family...I'm not going to pretend to know a lot about the disease, but I know that if you don't take care of yourself—I need to correct things—or if it runs in the family. And it does a little bit in my family—but again, it can be pretty bad. It's a nasty disease. I know that it's one that, apparently, is hard to control. | No, I don't think that—no [understanding didn't change]. |

*Participant ID is unknown to everyone outside the research team.

Key:

| | | | |
|---|---|---|---|
| Had personal experience or understanding (pre) | Maybe knows more (post) | Knows more or has decent understanding (post) | Doesn't know more (post) |

| Pt | Hypertension<br>*(What was your understanding of hypertension/high blood pressure before/after this project?)* | |
|---|---|---|
| | **Pre** | **Post** |
| 2 | I really didn't know anything until after the study. | I really still don't have an understanding of it. |
| 3 | Male: Hypertension?<br>Moderator: Um-hmm, high blood pressure.<br>Male: High blood pressure?<br>Moderator: Yeah.<br>Male: What was my understanding? I'm not very good at reading those. My [spouse]'s pretty good at that. [they're] the one that kind of looked at it. | Male: What was my understanding? I'm not very good at reading those. My [spouse]'s pretty good at that. [they're] the one that kind of looked at it.<br>Moderator: Was that information included in your result letter also?<br>Male: Yes.<br>Moderator: Did you understand that portion of the letter?<br>Male: Yes. |
| 4 | I know you don't want it [hypertension]. | For me, I know mine is really low. If I remember correctly, [my spouse], theirs was okay too. We just keep on trying to live an active healthy lifestyle. |
| 6 | Like I said, it's always been something that I've dealt with, with people I work with, also with family. So it's…nothing new. | Like I said, it's always been something that I've dealt with, with people I work with, also with family. So it's…nothing new. |
| 7 | I had an understanding of it. | I'm trying to think if your letter has something that went over hypertension. I think you could explain hypertension more. I mean, I might have an understanding of it, but somebody else might not understand—like maybe give like a blood pressure reading of what hypertension is and how it affects people. |
| 14 | It's not good for you, and I fully understand high blood pressure and hypertension. I understand it. It can kill you, and I need to pay more attention now. | I need to pay more attention now. |
| 16 | I would say it's low, my understanding. I mean, I know the importance of it, but… | Yeah, I feel like I know more, I think, with it being explained a little bit more and then actually doing the blood pressure and everything; it kind of helps out. |
| 27 | It's a killer...Nobody wants to have high blood pressure, especially people who have had heart issues like me. That's not good, because your heart, depending on what it is, is working more and you're more susceptible to strokes and—well, heart attacks has more to do with plaque and things like that, cholesterol. | It wasn't an issue for me, because I don't have high blood pressure. Even with all the heart issues I've had, my heart is actually very strong. It's probably because of working physically for all those years and outside of what I did, being so active. |
| 35 | It's not good. (Laughter.) I haven't had high blood pressure. | After participating, do you have any new information that you've learned about high blood pressure?<br>Male: No. |
| 39 | High blood pressure was a lot of stress, but now it's not so much the stress, that I understand it has a lot to do with a lot of other things. | High blood pressure was a lot of stress, but now it's not so much the stress, that I understand it has a lot to do with a lot of other things. |
| 41 | I have a good understanding of hypertension. | That it's a condition that needs to be looked at, that there are diets. There's a diet that you can use. |
| 54 | My [spouse] has high blood pressure, so I know what effects that can do and what it does to you. I have to watch [them] and be careful what he eats or what he cooks with, especially if I'm not there. Especially the goodies, boy. "Did you have ice cream?" "No." (Laughter.) Yeah, that I do know, because I'm around it all the time. | |
| 57 | Male: What's hypertension?<br>Moderator: Hypertension is like high blood pressure.<br>Male: They checked my blood pressure and it was | The last visit that I had, the guy said that having your blood checked and it was high, it could cause a stroke or a heart attack. At my age I think it's a good idea to check it at least two or three times a week… |

*Participant ID is unknown to everyone outside the research team.

Key:

| Had personal experience or understanding (pre) | Maybe knows more (post) | Knows more or has decent understanding (post) | Doesn't know more (post) |
|---|---|---|---|

| Pt | Hypertension<br>*(What was your understanding of hypertension/high blood pressure before/after this project?)* | |
|---|---|---|
| | **Pre** | **Post** |
| | okay. I feel okay. I check my blood pressure at least two or three times a week. | |
| 67 | I'm on medication for hypertension. | I don't know if it had anything to do with the study. I know that they reduced my medication for hypertension, but I don't think it had anything to do with the study that I felt better or worse, but it was too much, the dosage was too much for me, so they cut it in half, and I'm fine. |
| 68 | That's high blood pressure?...Aware of it because my family, some of my family members have high blood pressure, but I don't—not that I know of. | Moderator: Did it change at all after receiving your results letter?<br>Male: No. |
| 71 | | A little more, not that much more. I couldn't name stuff off right now because, like I said, I read that letter, except I can't remember what it was...If you'd have called me right after I received the letter, I could probably tell you everything you need to know, but like I said, it's been a while. |
| 76 | All I knew was your risk for probably a heart attack or stuff like that. That's about all. That's all I know now too. | All I knew was your risk for probably a heart attack or stuff like that. That's about all. That's all I know now too. |
| 90 | Female: I do know that it can kill you and it probably is a disease, right? How does one fix that? They have to get educated. They probably have to stay calm, try to find ways to be at peace with themselves and not get upset and eat right. Whatever abuse that that person has done with their physical body, then they have to do the same thing that I'm trying to do right now.<br>Moderator: Before the study, was that your understanding of what had to happen to prevent high blood pressure, or were those things you learned after you got your results?<br>Female: I don't have high blood pressure, I don't think, because I'm not on medication for it and they did testing and everything, or do I? No, okay. I know my [spouse], [they have] hypertension and so it scares me for [them], but it's [their] life too, and [they've] lived to this point, and [they're in their 60s]. How do change a person that has not had the discipline or the education? How do you change them after all that time? If [they do] discipline [themselves] and do what [they're] supposed to, how much life is it going to prolong for [them]? That's the thing. Parents should start teaching their kids when they're little the severity. It's just life. People are going to do what they want to do. That's what it comes down to. | |
| 110 | Hypertension was what I got when I was in boot camp, (laughter) almost getting a heart attack. Just totally zonked out, breathing hard, chest pains, nervousness, sweating, dizziness. That was my view of hypertension. But later on, I really began to understand the clinical view of hypertension. It goes along hand in hand with the heart a lot and with what you eat and what you put in your body. | But later on, I really began to understand the clinical view of hypertension. It goes along hand in hand with the heart a lot and with what you eat and what you put in your body.<br>Moderator: The more clinical understanding, is that something that happened after participating in the study?<br>Male: Yes. |

*Participant ID is unknown to everyone outside the research team.

Key:

| Had personal experience or understanding (pre) | Maybe knows more (post) | Knows more or has decent understanding (post) | Doesn't know more (post) |
|---|---|---|---|

| Pt | Hypertension<br>*(What was your understanding of hypertension/high blood pressure before/after this project?)* | |
|---|---|---|
| | **Pre** | **Post** |
| 116 | I don't know. (Laughter.) Hypertension would be like you're very hyper and stuff like that. You get nervous, all that stuff. | Moderator: After receiving the results letter, what's your understanding now?<br>Female: How do I understand it?<br>Moderator: Yeah, I'm just trying to ask about how your understanding might have changed before and after the study.<br>Female: Yeah, it would change. I would understand more what it's about, how to go about it and everything, how to help myself, somebody like that can work with me or something. |
| 117 | Bad eating, weight, diabetes causes high blood pressure, and it's not good. | Same understanding. Just what to do about it now...You take medication, exercise, all that stuff that I'm not doing. Eat better. Lower salt. I don't know if that's a thing. |
| 119 | It can cause a stroke. That's about all I knew then. | That you have to eat right, lower your salt. I drink a lot of water. That's about it. Just that, I guess. |
| 123 | I don't know anything about high blood pressure. I don't know how to answer that. | I don't know anything about high blood pressure. I don't know how to answer that. |
| 124 | I knew that it's very important to keep your high blood pressure down, your heart and everything else. I do understand that you have to maintain some kind of medication or whatever you need to do to keep it as close to normal as you can. | My understanding is that I've got to keep it up. Doing what I'm doing. |
| 127 | About the same. I didn't learn any more than what I already knew...I've been dealing with the hypertension also since before your study come around... When I had my first heart attack, back in '13. That's when I started finding out my hypertension was causing me a problem. | It's about the same. It was the same because I've been dealing with the hypertension also since before your study come around. |
| 131 | I didn't think of anything with health. I didn't think of even looking at or trying to get to know, so I didn't have any idea on blood pressure. | I don't want to get high blood pressure. To keep it under control. |
| 132 | High blood pressure? Is that, like, more having to do with what you eat and stuff? Yeah, that's basically all I know. | I guess, the same. |
| 141 | High blood pressure is due to stress and stuff, so it's good to stay within your normal range. It could lead to like—I don't know—maybe anything bad. I mean, it's just not good. That's all I know. | High blood pressure is due to stress and stuff, so it's good to stay within your normal range. It could lead to like—I don't know—maybe anything bad. I mean, it's just not good. That's all I know. |
| 145 | I didn't know nothing about high blood pressure. | I suppose I have high blood pressure. |
| 157 | It's almost something behavioral, I think. I think it has something to do with your blood pressure. I think—I'm guessing. | My understanding now there's actually stages of your blood pressure that can go like that. Doing the kidney research, you can find that out through that. That's something I learned afterwards. |
| 166 | Yes, it's only when your blood pressure is very, very high. Only when your blood pressure is normal and high. My blood pressure is normal, so they got me off the blood pressure pills. Physical, I don't know. I have to check my blood pressure, which runs high, which is low, and mark it down. Other than that, I know when my sugar is high and I don't feel good, I feel like I don't remember stuff, like, "Oh, my God. What did I do?" So I know what's going on, I have to count to ten | Do you feel like your idea of what high blood pressure was changed after taking part in the kidney study?<br>Female: No. My blood pressure—I don't know if my blood pressure changed or not. I never noticed. I need to ask my [sibling] that question, if your blood pressure changed. I think it stays normal or high. I do not know. |

*Participant ID is unknown to everyone outside the research team.
Key:

| Had personal experience or understanding (pre) | Maybe knows more (post) | Knows more or has decent understanding (post) | Doesn't know more (post) |
|---|---|---|---|

| Pt | Hypertension<br>*(What was your understanding of hypertension/high blood pressure before/after this project?)* | |
|---|---|---|
| | **Pre** | **Post** |
| | to calm myself down, and then my sugar will start going down. | |
| 194 | | I didn't know it could kill. It kills a lot of people. It's a problem. What I didn't realize was almost everyone has hypertension, it seems like.<br>Moderator: Those were things that you learned about after the study? Was it after the study, or was it after the results letter?<br>Male: During and after, yeah.<br>Moderator: Currently, after getting that letter, what is your understanding of hypertension?<br>Male: It can cause—along with what I read on hypertension, because I have it, that it's a silent killer. You could be alright one day, and run into trouble if you prolong treatment for it. |
| 195 | Moderator: What was your understanding of hypertension before the study?<br>Male: I didn't, and I don't know that I have a good feel for it now. | Moderator: What was your understanding of hypertension before the study?<br>Male: I didn't, and I don't know that I have a good feel for it now.<br>Moderator: After receiving the results letter, what would you say your understanding was of it?<br>Male: I don't remember that portion.<br>Moderator: You were saying that, before or even now, you feel like you don't have a great handle on what it is or just, in general, information on it?<br>Male: Yeah, right now, I'm drawing a blank on hypertension. |

*Participant ID is unknown to everyone outside the research team.
Key:

| | | | |
|---|---|---|---|
| Had personal experience or understanding (pre) | Maybe knows more (post) | Knows more or has decent understanding (post) | Doesn't know more (post) |

| Pt | Diabetes<br>*(What was your understanding of diabetes before/after this project?)* | |
|---|---|---|
| | **Pre** | **Post** |
| 2 | Just knowing that their blood sugar was low and you feel faint. That's all. | There's a real strict diet to it and just that you could lose body limbs and lose your kidney and eyesight. And it's real poisonous. |
| 3 | Sugar diabetes? Well, my understanding of diabetes is that there's a problem with the blood sugar level, which we can get at any time, especially as we get older, so it's just good to participate. | I think it's all good. We have a pretty good idea. My [spouse] likes keeping up with the stuff, so [they] keeps me posted. |
| 4 | I know it's a slow death, yes. | We have to be more mindful of our diet, really, is what it comes down to. |
| 6 | Like I said, it's always been something that I've dealt with, with people I work with, also with family. So it's…nothing new. | Like I said, it's always been something that I've dealt with, with people I work with, also with family. So it's…nothing new. |
| 7 | [Works at dialysis clinic.] I don't know what the letter has on there, like what causes it, like high blood pressure or diabetes. Maybe you guys could explain what causes it and how it can affect your health. | Yeah, but I don't think some people get it. Like, the A1c on there, I don't think people understand like if a high A1c, your sugar is high, like how their diet contributes to the diabetes and how it damages their kidneys. I don't think some people get that it damages their kidneys. |
| 14 | Again, it's something that can kill you. If you don't regulate it and you don't take care of it, it's something that will definitely kill you. It took my biological [parent]'s life, so it does concern me. | I'll be honest with you. I mean, our history goes back so long that, what is there to learn? It's hereditary for us, and it's a bad disease. |
| 16 | You know, it's still kind of low. I know people who have it. I don't have anybody with it in the family that has it, but I can see the importance. | You know, it's still kind of low. I know people who have it. I don't have anybody with it in the family that has it, but I can see the importance. |
| 27 | Diabetes, I don't think I knew that much about, except for people shouldn't eat a lot of sugar, but also, a contradiction was they should carry a candy bar with them…Because your sugar levels, you're supposed to monitor them, and if it gets out of whack, a candy bar can bring you back if you go into shock, because I know people who have kidney disease to some degree in the area, and that's one of the things. Diet is also key. It seems like most people like us when you get older, you get set in your ways, and they're just like, "Well, okay," but when you have a lot of things like myself, hey, I want to be here as long as I can. | No, it was still pretty much the same. I knew pretty much that I wasn't in that category, so I didn't—I'm not a person that would research it or look into it. I was just like, "Oh, that's good news for one thing." |
| 35 | It's a big problem in this area, especially, and there are cures. There are things that we can work on within ourselves, diet and exercise, etc. | Moderator: Has your understanding changed at all with this project?<br>Male: No. |
| 39 | Diabetes was when you had a lot of sugar. That's the way I thought it was, when you eat too much sugar, but it's not. It has a lot to do with your triglycerides, your—all these other fancy words that I can't say, because I've learned all those, believe me. | That's the way I thought it was, when you eat too much sugar, but it's not. It has a lot to do with your triglycerides, your—all these other fancy words that I can't say, because I've learned all those, believe me…I'm learning, because I just got diagnosed when I was fatter, and now that I'm losing it, my numbers are coming down. I'm learning about diabetes at the same time. |
| 41 | I think I understand diabetes fairly well. | Moderator: Did that change after participating in this study?<br>Female: No. |
| 54 | I've had a lot of information because of one of my coworkers is diabetic and that's how I learned and some of my elders I know they have it, too, so I have | You've just got to—it's a really bad disease. I've seen a lot of people, what it does to them. I understand it better than I did before, let's say, maybe eight years |

*Participant ID is unknown to everyone outside the research team.

Key:

| Had personal experience or understanding (pre) | Maybe knows more (post) | Knows more or has decent understanding (post) | Doesn't know more (post) |
|---|---|---|---|

| | | |
|---|---|---|
| | a good understanding of the information and what it does to you. They've got to be careful of what they eat. They have to do insulin and all this other stuff, so yeah, that's crazy. I thank God that I don't have to do that, but I feel bad for the people that do. | ago. I have a better understanding of it now than I did back then because of the people I'm surrounded by. |
| 57 | Nothing. Just what people talk about when they come out of dialysis. That's about it. That's about all I know. They have to go to dialysis because they're diabetics and kidney failure and things like that. I don't know very much about it. | Moderator: After getting your letter, the results letter, did you learn anything new about diabetes?<br>Male: No. |
| 67 | I had a thorough understanding of diabetes, because again, I'm type 1, and it started at a very early age. I was [a teenager] at the time. You learn to deal with it. There's not too many people that have survived on insulin for as long as I have, so I'm fine with it. It's part of life. As far as I'm concerned, it's no different than taking a shower and brushing my teeth. I just deal with it. | I had a thorough understanding of diabetes, because again, I'm type 1, and it started at a very early age. I was [a teenager] at the time. You learn to deal with it. There's not too many people that have survived on insulin for as long as I have, so I'm fine with it. It's part of life. As far as I'm concerned, it's no different than taking a shower and brushing my teeth. I just deal with it. |
| 68 | That's what got me going on the testing, and then, after you guys gave me the results and my doctor got together with me, my kidneys were taking a beating because of pre-diabetes… Now, I have to take care of that. | That's what got me going on the testing, and then, after you guys gave me the results and my doctor got together with me, my kidneys were taking a beating because of pre-diabetes… Now, I have to take care of that. |
| 71 | That it's a deadly disease. You could die from it. You can lose limbs from it. You could go blind from it. | Just to maintain a healthy diet, watch what you eat. Like I said, part of the reason for diabetes is salt, whatever, sugar, just blah blah, blah, bad stuff. Control it. Don't let it control you. |
| 76 | …when I started getting all this, gaining more weight and getting all these problems and everything. The diabetes, all I knew, that if you weren't careful and control your diabetes, they could amputate your joints, your feet, or whatever. That's pretty much about it. [I knew] to exercise and lose weight, cut down on the food you eat, not so much, especially sugary stuff and things that we like to eat. | It's pretty much the same. It's just a thing that, like I said, making up your mind to do something about it, which is, I think, most of our problems, that we don't take it so serious. We actually see somebody going through all their problems. |
| 90 | I know that my [sibling]-in-law had diabetes really, really bad. They have to monitor their intake, or they could die. The body parts, they get amputated. [they] didn't have any of [their] body parts amputated, but it goes down to the way we eat, it really does, and how we take care of our bodies. Everything needs to be taken care of just like if you have a machine you have to oil it; you have to put gas in it; you have to take care of it. That's how our bodies are. We have to take care of our bodies, and a lot of people don't. That's why there are all these medical issues. Then again, it could be hereditary as well, so it has to do with the DNA. | I know that my [sibling]-in-law had diabetes really, really bad. They have to monitor their intake, or they could die. The body parts, they get amputated. [they] didn't have any of [their] body parts amputated, but it goes down to the way we eat, it really does, and how we take care of our bodies. Everything needs to be taken care of just like if you have a machine you have to oil it; you have to put gas in it; you have to take care of it. That's how our bodies are. We have to take care of our bodies, and a lot of people don't. That's why there are all these medical issues. Then again, it could be hereditary as well, so it has to do with the DNA. |
| 110 | I thought only older people that indulge in sweets were susceptible to such. I was diagnosed when I was in my late thirties. I went from type 1 or type 2 in just a matter of a couple of years. I just totally had no idea that it was something that will happen to somebody in their thirties, or early forties for that matter. I thought it was somebody that just sat there and indulged in chocolate cake all day and didn't work out, somebody old, just sat there, a couch-potato-type thing, but I'm active. I was a carpenter for a while. I | It can be controlled, and it can progress. It could be one's demise, but it's something you got to live with and something you got to treat and keep in check, not only with exercise and watching what you eat, but in terms of medications, if need be. I'm on metformin now. I get the nerve pains, and my sight's going. It's hit me in all directions, and I see it as very dangerous disease. |

*Participant ID is unknown to everyone outside the research team.

Key:

| Had personal experience or understanding (pre) | Maybe knows more (post) | Knows more or has decent understanding (post) | Doesn't know more (post) |
|---|---|---|---|

| | | |
|---|---|---|
| | walk a lot. I thought I was going to be the last candidate to be designated as a diabetic, but that all changed one day when I ended up in the hospital. They told me, "Your blood sugar is—you're about to go into a coma. You're in your 300s." I'm obese, a fat layer, but I never thought I would be in that predicament. | |
| 116 | That it has to do with your kidneys and how obesity [sic] you are and the food that you eat, things like that. You need to take medication, if you have to. | I would understand it like we need to take care of ourselves, what we eat, how we drink water and stuff like that. |
| 117 | I have a lot more understanding of diabetes even before the project, not any more or any less understanding since before this. I kind of got my reality check when I got thrown in the hospital a way back. That was my real reality check on that. | I have a lot more understanding of diabetes even before the project, not any more or any less understanding since before this. I kind of got my reality check when I got thrown in the hospital a way back. That was my real reality check on that. |
| 119 | I've had diabetes for some time. I know when I get a low blood sugar, it just drains me. I've got to be careful and not let that happen. Just general diabetes information. My family has had it for years. I've heard about different things off and on, but nothing formal, no formal teaching or anything like that. | I've had diabetes for some time. I know when I get a low blood sugar, it just drains me. I've got to be careful and not let that happen. Just general diabetes information. My family has had it for years. I've heard about different things off and on, but nothing formal, no formal teaching or anything like that. |
| 123 | Diabetes is—I'm not sure about that either. I know what it is, in a sense. I know it's bad, but I don't know that much about it. | Diabetes is—I'm not sure about that either. I know what it is, in a sense. I know it's bad, but I don't know that much about it. |
| 124 | Diabetes, like I said, I already had it for about 20 years. I follow the instructions that usually my primary doctor tells me. I do what they tell me to do. I try to do the prescriptions that they give me or cut down on this or up on the other. That's about it. | I know that you've got to stay on top of it, on your medications and all that. That's about it. |
| 127 | …my family members have had diabetes issues, so I've learned about diabetes since I was a kid. | [The letter] didn't help me any. As far as my understanding, I already knew. There's not much more. I know quite a bit about diabetes, and I know quite a bit about kidney disease, since before this study, because [of my family]. I didn't really learn anything. |
| 131 | I didn't understand diabetes, because we have diabetics in my family, so I was just scared of diabetes. | That you get really sick. When you become a diabetic, you get really sick off of it. You can keep doing what you're doing to keep you from getting sicker, but in the long-run it's not good. I've seen my family go from, you know, all the way down. That's a good reason, too, that I needed to change my life, because of diabetes in my family. It's on both sides. |
| 132 | I know this one. I get nervous when I talk. I'm sorry. It's having to do with your blood, your sugars and stuff. | I guess, the same, just that it's a lot more worse than I thought it was. It's a bigger deal than I thought. |
| 141 | Diabetes could be genetically passed on through family members, or it could be contracted through your eating habits, your lifestyle, and it's just the intake of whatever you're eating, I guess. It could lead to just the malnutrition from whatever you're taking in. It's just not good for you. Changing your ways is way much better than getting that, and it could make your kidneys fail from it also. | My understanding now is pretty much the opposite of whatever I just explained. It's more of a positive side of thinking about it, even educating my little ones about the awful things that you should stay away from like sugar and fats and all of that other stuff. |
| 145 | Diabetes? I don't know. I don't really know anything about these questions, I guess. | Moderator: Now, has your understanding of diabetes increased? Decreased? Stayed the same?<br>Male: I don't have diabetes.<br>Moderator: But have you learned more about |

*Participant ID is unknown to everyone outside the research team.

Key:

| | | | |
|---|---|---|---|
| Had personal experience or understanding (pre) | Maybe knows more (post) | Knows more or has decent understanding (post) | Doesn't know more (post) |

| | | |
|---|---|---|
| | | diabetes?<br>Male: No. |
| 157 | Your body had too much sugar. That's kind of all I knew, that your body had too much sugar and you get diabetes from it. | Now I know there's—like I was saying, there's actually ways you can try to prevent that or try to better your health for that, just in case, like I said before, if your body ever goes in that route or your health ever goes in that route. I think it's a better understanding now with the kidney research and stuff, to kind of give people a heads up, just in case. |
| 166 | Diabetes is terrible for me. My sugar is really high. Usually it's high and low. My sugar gets high in the morning. I have to give myself 45 units of shots. I hate shots sometimes, to give myself, but I already got used to it. It's just sad because I'm diabetic because it runs in the family. | Actually, it has changed. It has been going on 100. I cut down on eating certain things. I cut down on eating lard. I'm eating a different lard for diabetics because I googled it. I cut down on Diet Cokes. I cut down on all the diet stuff that I had to cut down. I eat nothing but salads. |
| 194 | It seemed like I didn't know too many people that it affected, but now I see it…I lost quite a few distant relatives. Me, myself, I lost two [sibling]s to it. | It seemed like I didn't know too many people that it affected, but now I see it. It seems like more people have it…It's very devastating to people. You'll see them healthy. You remember them being healthy in their younger years. Then, when you see them get full-blown diabetes, it's terrible. It's terrible. They're no longer the same people that you knew before. They can't live a normal life…A lot of them, when they go in for the cleansing, when they have to go in for dialysis or something, it takes up most of their day. Then, by the time they get home, they're too exhausted to go out. You don't see them in the community. You would see them in town, at grocery stores or basketball games or whatever.<br>Moderator: This understanding of diabetes now and what you've been talking about, did that come after receiving the results letter?<br>Male: It had some to do with it, I think. |
| 195 | There's a little bit of diabetes in our family, so I've seen it there. Then I've seen where it's really had a negative effect on people I know, the customers that come in. When you lose a leg, lose an arm, lose whatever. Diabetes just eats you up. | I don't think the letter changed my understanding of diabetes. |

*Participant ID is unknown to everyone outside the research team.

Key:

| | | | |
|---|---|---|---|
| Had personal experience or understanding (pre) | Maybe knows more (post) | Knows more or has decent understanding (post) | Doesn't know more (post) |
