## Appendix 1 for "New Directions from the COMPASS Study: Participation and Communication in Rural Health Research"

**Appendix 2.** Expanded codebook with additional quotes and number and percentage of participants with quotes related to each code, split between main themes of Community and COMPASS.

| Codes and Sub-codes | Description | Example Quotes | Participants N | % |
|---|---|---|---|---|
| **Community** | | | | |
| **1. Health problems** | Health problems in the Cuba community. Includes barriers to healthcare. | • *…a friend of mine [had] kidney surgery. They removed one kidney Monday. I know of one, two, three, four, five—eight or so individuals that have kidney troubles, and probably ten or so that have diabetes.*<br>• *…When you see them get full-blown diabetes, it's terrible. It's terrible. They're no longer the same people that you knew before. They can't live a normal life. They have to go in for treatments. You don't see them very often anymore because that's mainly what they do…Then, when you do see them again, they look terrible. Then, before you know it, you hear that they've passed on, and it's sad.*<br>• *Like, people on drinks and stuff, a lot of people are dying from it. I mean, that's pretty much what's going on around the community. That's what I notice, like kidney failure and all that.*<br>• *Just really not good food decisions, not having money to make good food decisions, because really good food is slightly expensive…It's a very poor community.* | 33* | 100.0% |
| **2. Resources** | Resources in community or absence of. | • *What we need is more hospitals here in Cuba…Why do people have to go all the way to Albuquerque when we have big buildings here?*<br>• *…Thinking about drinking that water my entire life here, I'm pretty sure that I put my kidneys through a heck of a lot, rather than just with booze. I'm not the only one. There's these folks around here that can't afford bottled water. They rely on tap water for drinking, cooking, and cleaning. A lot of that, I think, goes hand in hand with kidney disease that's spreading out in rural communities because the water is not up to par and is not put in check and monitored like it's supposed to.* | 15 | 45.5% |
| **COMPASS** | | | | |
| **1. Considering participation** | | | 20* | 60.6% |
| a. Awareness/ understanding | Reason for participating is to raise awareness of or increase understanding of kidney-related health issues. | • *Just seeing that something could be wrong with my kidneys…so I can fix it before it gets too late, or change something before it gets worse, because I'm scared of not being able to live that much longer. I have kids.*<br>• *I have a lot of issues that run in my family, so I was concerned for my health. That's why I participated.* | 10 | 30.3% |
| b. Help community/ contribute to science | Reason for participating is wanting to help community or contribute to science. | • *…contribution to the future of our kids…I'd probably want to do it them and my relatives and my neighborhood, my neighbors, and community for generations to come. Because what we have today and in my lifetime, our parents and grandparents, they struggled. They went through a lot. They* | 9 | 27.3% |

| | | | | |
|---|---|---|---|---|
| | | *contributed so that we have the medications and medicines and knowledge that we have that kept me alive.*<br>• *I participate because I'm curious about myself, but it's something that I think is good for the community and try to get people involved in.*<br>• *I just wanted to be as helpful as I could be to the study. I think it's important to do research on things, on kidney failure and stuff, so we can figure out more of the problem.* | | |
| c. Participation concerns | Concerns, discomfort, fear specific to participation in the study, not about personal health. | • *Probably just what you need, like what do you do, like urine samples and drawing blood and stuff…It didn't worry me. I was just kind of curious. I wanted to know what you need to be taking to get your results for that…just a little explanation.*<br>• *I had never done [a study] before, so I guess it was just kind of like curiosity, but so far, it's fine.*<br>• *I don't really like needles, but it wasn't too often [that I needed to give blood]…I was kind of nervous, but I didn't freak out.* | 8 | 24.2% |
| **2. Experience** | | | 33* | 100.0% |
| a. Results letter | Comments about the results letters. | • *…I recall getting good information that was interesting, but it's been so long that I just don't remember what was specifically on there.* | 33* | 100.0% |
| • Lifestyle changes | Lifestyle changes they made (or didn't make) because of results letter. | • *It gave me an idea of where my health is and everything. It kind of helped me a little bit better to start taking care of my body and my kidneys and things like that more.*<br>• *Yeah, I drink a lot more water now. I try to watch what I eat, not too much fatty foods. I've lost like 25 pounds now.*<br>• *I think, at that time, we were drinking a little bit more, so we kind of just stopped drinking, just once in a blue moon.*<br>• *…I stopped exercising…I kind of lost interest in it [thinking about lifestyle changes].* | 26 | 78.8% |
| • Health literacy | Any mention of having trouble understanding what was in their results letter (or not). | • *Everything was easy to read. The only thing I remember is I understood it. I was happy with it. I didn't have to call a doctor to explain it to me. That's all that really mattered to me.*<br>• *"I think it'd be more informative, in the sense that the numbers were a little bit too technical, I guess is the word I should say. For an uneducated person, I could see that it would be very difficult to read or understand…less educated, not uneducated"*<br>• *Not being in the medical field, some of the things we didn't understand…I know you wrote down what was acceptable and not acceptable and where we were at, but some of the terms, I didn't know what they meant...* | 18 | 54.5% |
| • Third party | If they took the results letter to someone for any reason. | • *…There were a few numbers that we weren't too sure about. We have a friend who's a nurse, and so we let her look over what the study was.*<br>• *I think my sodium level was out of range, and I Googled it, what's normal range…* | 20 | 60.6% |

| | | | | |
|---|---|---|---|---|
| | | • *It was right on, because I took it to my doctor because I have issues, and it was right on point. He was very happy with the letter that you guys sent.* | | |
| b. Value | Value stemming from the experience of being in the study. | • *…You can tell a lot more from the blood screening than you can from just going to the doctor and saying, 'I'm sick,' because they just treat the symptoms, and they don't really get down to what's causing it.*<br>• *I don't go to the doc that often…I haven't had a physical in I don't know how long, maybe about 25 years or so. I'd like to get an idea of what my insides might be looking like…*<br>• *It was nice to find out where your ratings were, and even with it being where it needed to be, it was comforting to find out where I was.*<br>• *It lets you know if your kidneys are functioning, if there's something going on. The information is good to have, especially if you don't know. If there is something wrong, at least you'll be aware, and you're not just going through your years wondering, and then at the end, when it gets really bad, then you're like, "Oh, I didn't know."* | 27 | 81.8% |
| c. Health education | Knowledge of chronic kidney disease, hypertension, and diabetes. | See Table 4 and Appendix 3 for example quotes and more information about this code. | 33 | 100.0% |
| **3. Recommendations** | | | 33* | 100.0% |
| a. Results letter | How to improve the results letter. | • *I know that some people had a hard time reading their results, so maybe if you guys would have put what is maybe normal and abnormal. I think it's kind of on there, but I think the older people had a harder time understanding it…I don't think people understand if a high A1c, your sugar is high, like how their diet contributes to the diabetes and how it damages their kidneys.* | 9 | 27.3% |
| b. Study | Recommendations for how to improve the study, generally. | • *Maybe just have more classes out where we live and letting people know, like, nutrition that they could go by daily…*<br>• *For us elders…If they could maybe give us a call back or even another letter just to inform us say, "You know what? We haven't received a response," or something to that effect would be nice…[And] just more information maybe in writing would be good.*<br>• *I guess it would be nice to have someone tell me what kidney disease is all about and what to expect if I get it and what the symptoms are and that sort of thing, because I don't know.* | 9 | 27.3% |
| c. Recruitment | Considerations to improve recruitment. Includes how they found out about the study (what works). | • *My daughter works at the clinic, so she's the one who informed us, so word of mouth, but then we did find out about it, so we did refer a lot of our friends to it.*<br>• *In Cuba, sit in front of the post office…everybody goes to check their mail in a small town. You do it every single day religiously.* | 33 | 100.0% |

| | | | | |
|---|---|---|---|---|
| | | • *A lot of people don't like new things or change, just from what I've seen. When you try to put something new, it's kind of like a warning sign. Usually, once you get some people out there talking about it and doing it, sometimes you can develop interest in people.*<br>• *If you guys have classes talking about the kidneys, how it does to people or how it makes people feel. That would be cool. For the little kids, too, where they could know what kidneys do for people, what makes them sick. They could take care of themselves and anything. You guys need to have a school, talk about kidneys more better that people could understand.*<br>• *The last time, we made flyers, and we passed them around. The thing is, where I work at, it's a different county. There are others who would like to participate, but they're in a different county…*<br>• *I just know that I had heard previously from community members that it would be easier for them to participate if there was a later clinic or on the weekend or something that they could do, because it was hard for them to get out for their lunch…* | | |
| **4. Dissemination^** | Whether participants are interested in seeing study publications, plus ideas to share with the community. | • *Might as well see what I contributed to…to see what the results were…If it says "Cuba area," it'd probably be worth reading what's happening in my area, as far as kidney and stuff like that.*<br>• *Not if I can't do anything about it. I'd rather you send them to somebody that might do something about it instead of to me. I can't do nothing about it…If you all find something in the water department troubles, in the air quality troubles here in this town or in the city of this town, hopefully you guys can let somebody else know about it that might be able to fix it.*<br>• *Probably letters. It makes it a lot more personal anymore. You can get emails or whatever, and it doesn't seem special or anything anymore. Today, when you get a letter or something, you pay attention.*<br>• *Maybe pass it to the folks here at the Cuba clinic. Maybe some sort of a little seminar or a little something that they could pass on to the public as a whole, or the visitors that they have on a daily basis.* | 9 | 27.3% |
| **5. Future participation** | If and why they would be interested in participating in future research. | • *Because I like to know what's going on with my body, especially now that I know a lot of different things and I would love to know what else I can do to help myself.*<br>• *You make a little bit of money for your blood. (Laughter.) No, it's just good to keep up with—make sure everything's good, especially as you get older.*<br>• *I think it's good for people to be informed. It's good to be helpful and not just be ignorant towards new findings.*<br>• *I think helping find causes or reasons why things happen is just beneficial to everybody.* | 33 | 100.0% |

| | |
|---|---|
| | • *Just trying to help us and the best way to advocate for my people, because that's all that there is, because pretty much all of the people that I know, they all passed away from liver cirrhosis.* |

**Aggregated and unduplicated number of participants for parent and sub-codes (e.g., if a person has one quote in a sub-code and none in the parent code, their n of 1 is captured in the parent code and in the sub-code; if a person has one quote in a parent code and another quote in the sub-code, they are not counted twice and the aggregate n=1).*

^Questions about dissemination were added late in the study and only asked of nine participants.
